# Rich Oleocanthal and Oleacein Extra Virgin Olive Oil and Inflammatory and Antioxidant Status in people with Obesity and Prediabetes. The APRIL Study: a Randomised, Controlled Crossover Study

**DOI:** 10.1101/2023.03.24.23287704

**Authors:** Ignacio Ruiz-Garcia, Rodolfo Ortíz-Flores, Rocío Badia, Aranzazu Garcia-Borrego, María García-Fernández, Estrella Lara, Elisa Martín-Montañez, Sara García-Serrano, Sergio Valdés, Montserrat Gonzalo, María-José Tapia-Guerrero, José-Carlos Fernández-García, Alicia Sánchez-García, Francisca Muñoz-Cobos, Miguel Calderón-Cid, Rajaa El-Bekay, María-Isabel Covas, Gemma Rojo-Martínez, Gabriel Olveira, Silvana-Yanina Romero-Zerbo, Francisco-Javier Bermudez-Silva

**Author notes:** Correspondence: FJBS,;. SYRZ. These authors contributed equally (Ignacio Ruiz-Garcia, Rodolfo Ortíz-Flores).

## Abstract

Oleocanthal and oleacein are olive oil phenolic compounds with well known anti-inflammatory and anti-oxidant properties. The main evidence, however, is provided by experimental studies. Few human clinical trials have examined the health benefits of olive oils rich in these polyphenols. Our aim was to assess the health properties of rich oleocanthal and oleacein extra virgin olive oil (EVOO), compared to those of common olive oil (OO), in people with prediabetes and obesity. This was a randomised, double-blind, crossover trial done in people aged 40-65 years with obesity (BMI 30-40 kg/m^2^) and prediabetes (HbA1c 5.7-6.4%). The intervention consisted in substituting for 1 month the oil used for food, both raw and cooked, by EVOO or OO. No hypocaloric diet or changes in physical activity were recommended. The primary outcome was the inflammatory status. Secondary outcomes were the oxidative status, body weight, metabolic status and lipid profile. An ANCOVA model adjusted for age, sex and treatment administration sequence was used for the statistical analysis. 91 patients were enrolled (33 men and 58 women) and finished the trial. A decrease in interferon-γ was observed after EVOO treatment, reaching inter-treatment differences (P=0.041). Total antioxidant status increased and lipid and organic hydroperoxides decreased after EVOO treatment, the changes reaching significance compared to OO treatment (P<0.05). Decreases in weight, BMI and blood glucose (p<0.05) were found after treatment with EVOO and not with OO. In conclusion, treatment with EVOO rich in oleocanthal and oleacein differentially improved oxidative and inflammatory status in people with obesity and prediabetes.

## 1. Introduction

In the 1960’s, the Seven Countries Study showed that mortality due to coronary heart diseases in the Mediterranean area was 2–3 times lower than in North Europe and the USA[1]. This finding was attributed to the Mediterranean diet (MedDiet) whose central feature is the utilization of olive oil as the main source of fat. Traditionally, the types of olive oil consumed have been virgin olive oil (VOO) and extra virgin olive oil (EVOO) which are obtained from the fruit of the olive tree (*Olea europaea* L.) solely by mechanical means, the latter displaying better physico–chemical and organoleptic characteristics [2]. However, nowadays the most consumed is common olive oil (OO), a mix of refined olive oil (a lipid matrix obtained from low quality VOO after an aggressive physical and chemical industrial processing) and a low percentage of VOO.

Together with the particular fatty acid composition of olive oil, rich in the monounsaturated oleic acid, VOO and EVOO has minor components (about 2% in weight) including phytosterols, tocopherols, and polyphenols, while OO has only traces[3]. During some time, the high content of monounsaturated fatty acids (MUFAs) was considered to be responsible for the protective effects of olive oil. However, now it is known that most of these benefits are also related to the minor components in the unsaponifiable fraction, particularly the phenolic compounds. Thus, olive polyphenols are considered to be responsible for some of the recognized pharmacological properties of the olive tree (anti-atherogenic, antihepatotoxic, hypoglycemic, anti-inflammatory,anti-oxidant, antitumoral, antiviral, analgesic, purgative and immunomodulatory activities), together with the protection against aging-associated neurodegeneration[4].

Nutritional interventions using olive oils rich in anti-inflammatory polyphenols could be of special value for people with obesity and/or at risk of developing T2D given the important role that inflammation plays in insulin resistance and diabetes pathogenesis[5,6]. In fact, a meta-analysis of 30 human intervention studies with olive oil showed overall the amelioration of the antioxidant and inflammatory status of the subjects, the beneficial effects being more pronounced in subjects with an established metabolic syndrome or other chronic conditions/diseases[7].

In recent years oleocanthal and oleacein has received much scientific interest due to their biological properties, particularly their ability to modulate inflammation, oxidative stress and cell proliferation [8,9]. Oleocanthal was also identified as the responsible for the throat irritant or stinging sensation produced by some EVOOs[10], similar to that caused by the non-steroidal anti-inflammatory drug ibuprofen[8,11].

To the best of our knowledge, there are only three clinical studies having examined the health benefits of rich oleocanthal and/or oleacein olive oils. One was performed with only 9 healthy individuals and it examined the effects of two similar virgin olive oils but with differences in their phenolic content, one rich and the other poor in oleocanthal, on platelet aggregation[12]. Results suggested an acute anti-platelet effects in healthy men[12]. Another pilot study tested an intervention with high oleocanthal and oleacein EVOO in 26 patients at early stage of chronic lymphocytic leukemia[13]. Authors concluded that it could be a promising dietary approach for the improvement of this disease[13]. The third study was performed in 23 subjects with metabolic syndrome and hepatic steatosis[14]. They were administered rich oleocanthal EVOO for two months and the effects were assessed before and after the intervention. The results showed that ingestion of EVOO with a high oleocanthal concentration had beneficial effects on metabolic parameters, inflammatory cytokines and abdominal fat distribution [14].

Our aim was to examine the health effects of a rich oleocanthal and oleacein EVOO, versus those of a common olive oil, in people with obesity and prediabetes. Given that inflammation and oxidative stress are intertwined processes which participate in the etiology and physiopathology of obesity and type 2 diabetes, we conducted a clinical trial in which the primary outcome was the modulation of inflammatory markers as assessed in a panel containing both anti-inflammatory and pro-inflammatory cytokines, and secondary endpoints were the oxidative status and relevant clinical parameters such as body weight, BMI and metabolic status (fasting glucose, insulinemia, HOMA-IR, HOMA2 IR, QUICKI, HOMA2%B, HOMA2%S, HbA1C and lipid profile).

## 2. Material & Methods

### 2.1 Study design

The APRIL (Aove in PRedIabetes) study was a randomized, crossover, double-blind, controlled clinical trial with 1:1 assignment performed between July 2016 and June 2019 at the Regional Hospital of Málaga–Biomedical Research Institute of Málaga (IBIMA), Málaga (Spain). People with obesity and prediabetes were randomly assigned to receive EVOO or OO during the first intervention period, 30 days, via a random number generator. EVOO and OO were provided in coded bottles, masked for both patients and researchers. After the first intervention period, a washout period of 15 days was left to eliminate the carryover effect, in which the subjects returned to the oils they used regularly before beginning the study. Subsequently, they were submitted to the second intervention period, again for 30 days, in which they consumed the other olive oil (OO for people initially treated with EVOO, and EVOO for those previously treated with OO). The general scheme of the intervention is shown in Figure 1.

**Figure 1.**
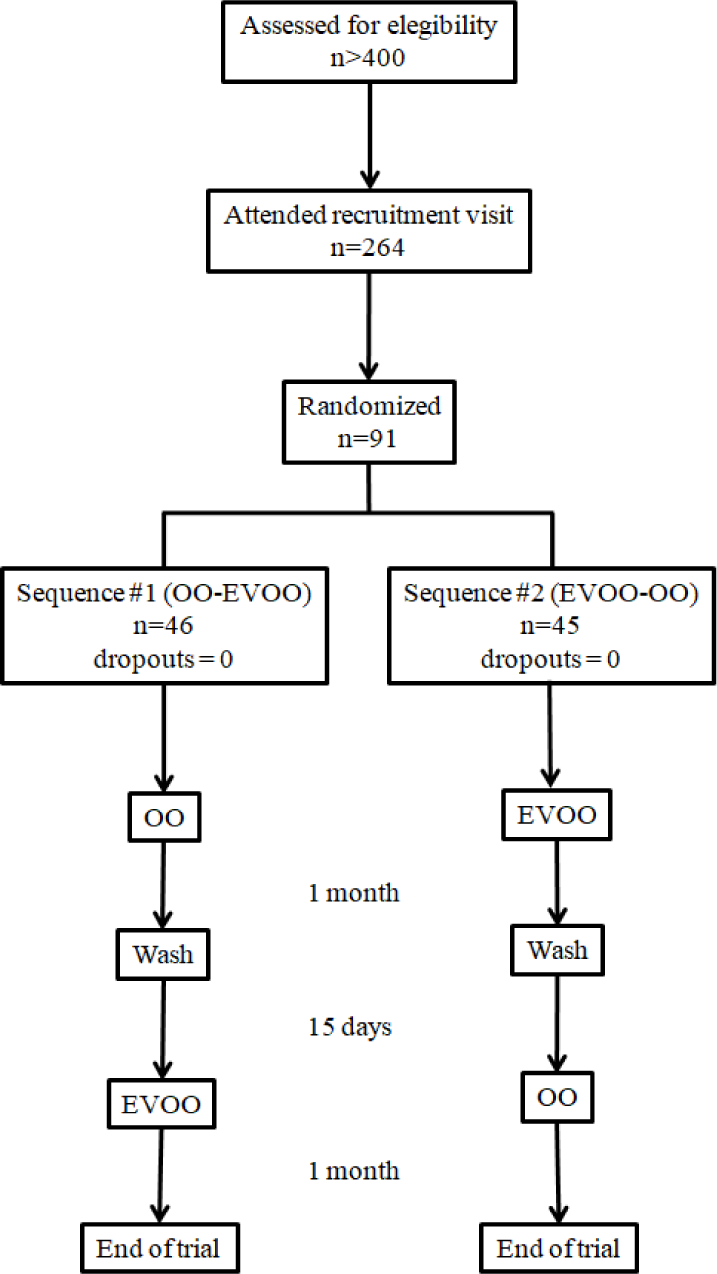
Flow diagram of the APRIL study.

### 2.2 Selection of olive oils and phenolic content analysis

Given the current evidence on the healthy properties of secoiridoids aglycons in olive oil such as oleocanthal and oleacein, an EVOO rich in these compounds was selected for the study, together with an OO with low levels of these aglycons but with similar levels of simple phenols such as tyrosol and hydroxytyrosol, also reported in the literature as beneficial polyphenols. The OO, consisting of a blend of refined olive oil and VOO fit for consumption as is (19). The phenolic composition of the two olive oils is shown in Table 1. Overall, EVOO contained 7-fold higher amount of total polyphenols than OO (508.4 vs 76.83mg/Kg), with secoiridoids constituting 93% of total polyphenols in EVOO (472.91 vs 66.11mg/Kg). Oleocanthal (69%) and Oleacein (21%), 428.31mg/Kg in total, constituted the main secoiridoids in EVOO.

**Table 1.** Concentration (mg/kg) of phenolic compounds in olive oils, common olive oil (OO), and extra virgin olive oil (EVOO).

| Class | Phenolic compound | OO | EVOO |
| --- | --- | --- | --- |
| Simple phenols | Hydroxytyrosol | $1.86 \pm 0.31$ | $1.75 \pm 0.85$ |
| | Tyrosol | $7.50 \pm 2.09$ | $6.84 \pm 2.83$ |
| | Hydroxytyrosol acetate | nd | $0.44 \pm 0.02$ |
| Secoiridoids | Oleacein | $9.22 \pm 1.02$ | $99.70 \pm 5.32$ |
| | Oleocanthal | $42.69 \pm 8.87$ | $328.61 \pm 15.69$ |
| | Oleuropein aglycon | $8.18 \pm 2.07$ | $25.79 \pm 3.44$ |
| | Ligstroside aglycon | $6.02 \pm 0.41$ | $18.81 \pm 2.60$ |
| Lignans | Pinoresinol | $0.22 \pm 0.05$ | $1.48 \pm 0.52$ |
| | 1-Acetoxypinoresinol | $1.00 \pm 0.01$ | $14.68 \pm 3.76$ |
| Flavonoids | Luteolin | $0.14 \pm 0.03$ | $1.89 \pm 0.47$ |
| | Apigenin | nd | $8.41 \pm 1.02$ |
| Sum of phenols | | $76.83 \pm 14.86$ | $508.40 \pm 34.52$ |

Not detected (nd). Values are expressed as mean ± SD (n=2).

The phenolic content was determined by liquid chromatography (HPLC) using the method recommended by the International Olive Council with modifications[15]. Briefly, a liquid/liquid phenolic extract was made with aqueous methanol, the extract was injected into HPLC where the phenols were separated on a C-18 reverse phase column with methanol-water gradient elution. Detection was performed with a diode detector (UV-DAD) and by mass spectrometry (HPLC-ESI-MS). Standard compounds hydroxytyrosol, tyrosol, hydroxytyrosol acetate, 4-methylcatechol, luteolin and apigenin were all purchased from Sigma-Aldrich (St Louis, MO). The dialdehydic form of elenolic acid linked to hydroxytyrosol -3,4-DHPEA-EDA (oleacein), dialdehydic form of elenolic acid linked to tyrosol -*p*-HPEA-EDA (oleocanthal), oleuropein aglycon -3,4-DHPEA-EA, ligstroside aglycon -*p*-HPEA-EA and pinoresinol, were all purchased from Phytolab (Germany). 1-acetoxypinoresinol was obtained from VOO in an analytic C-18 column and eluted with MeOH:H_2_O [15,16].

### 2.3 Participants and recruitment

Eligible participants were men and women (aged 40-65 years) with obesity (BMI: 30-40 kg/m^2^) and prediabetes (assessed as glycated haemoglobin 5.7-6.4). Exclusion criteria were previous diagnosis of diabetes mellitus, pregnancy, neoplasia, inflammatory diseases or being under treatment with anti-inflammatory drugs, women on hormone replacement therapy, and eating outside three or more meals (lunch or dinner) during the week. Candidates were selected from Pizarra’s study database (an epidemiological study of diabetes incidence previously done by our research group)[17], the obesity outpatient unit of our hospital and two primary care centers (El Palo and Alhaurin el Grande, Málaga). Subjects potentially meeting inclusion/exclusion criteria were invited to attend an examination visit at the hospital or health center in fasting condition and providing a sample of urine and faeces. In this visit the patients were informed on the nature and characteristics of the study and the inclusion/exclusion criteria were assessed. Those meeting the criteria and agreeing and signing the informed consent were enrolled in the study.

### 2.4 Intervention, data collection and sampling

The intervention consisted in substituting the main source of lipids in diets (in Spain it is generally a common olive oil) by the one assigned to each period. No specific amount of olive oil intake was indicated and patients were encouraged to maintain the same dietetic and lifestyle habits with the exception of changing the oil. No advices on reducing caloric intake or changing physical activity/sedentary behaviours were given. In order to secure that participants ingested the assigned olive oil even in meals needing home cooking, the amount of olive oil provided took into account the number of family members. Specifically, participants received 4 L/month (for families comprising up to 3 members) or 6 L/month (more than 3 members in the family). At baseline and after 30 days of intervention, information was collected using an interviewer-administered structured questionnaire, followed by a physical examination by a nurse (the same in all cases), who prior to the study had undergone a specific training course. Socio-demographical data were collected, as well as information on smoking, level of daily physical activity, leisure time sporting activity, alcohol intake and any changes in quantity or type of food in the last 6 months. For this latter purpose, a food frequency questionnaire was filled [18]. MedDiet adherence was assessed by using the validated brief 14-items questionnaire[19]. Physical activity was assessed by the International Physical Activity Questionnaire - Short Form (IPAQ)[20]. Anthropometric and clinical data (medical condition and use of medication) were also recorded as well as pulse and blood pressure. Waist circumference measurements were made on bare skin and hip circumference and weight measurements over underwear. Height was calculated with a stadiometer (Holtain Limited, Crymych, UK) and weight with a scale set to 0.1 kg (SECA 665, Hamburg, Germany). Body mass index (BMI) was calculated as weight ⁄ height (kg/m^2^). Pulse and blood pressure were measured by using a blood pressure monitor (Hem-703C; Omron, Barcelona, Spain) following instructions of manufacturer. Blood samples were withdrawn from the cubital vein and collected in sterile plastic tubes with a vacuum system. Plasma and serum were immediately obtained after centrifugation and stored at −80°C until analysis. Samples were stored in the Biobank of the Andalusian Public Health System (Regional Hospital of Málaga, Spain).

### 2.5 Direct and surrogate biochemical and metabolic determinations

Blood samples for biochemical determinations were immediately sent to the Laboratory of Analysis and Clinical Biochemistry of the Regional Hospital of Malaga, to be analyzed by hospital routine methods for glucose, insulin, HbA1c, 25-OH Vitamin D, TSH, C-reactive protein (CRP), both normal and high-sensitivity, as well as for other common biochemical markers such as urea, creatinine, uric acid, triglycerides, total cholesterol and HDL cholesterol. LDL cholesterol was estimated by the Friedewald calculation and Glomerular Filtration Rate (GFR) by the CKD-EPI equation. Insulin resistance was calculated by the homeostatic models HOMA-IR and HOMA2 IR index, as well as by the quantitative insulin sensitivity index (QUICKI). HOMA2 calculator was also used to estimate beta cell function (%B) and insulin sensitivity (%S). HOMA-IR was calculated as: (Fasting Insulin x Fasting Glucose)/405; QUICKI was calculated as 1 / ((log (fasting insulin μU/mL) + log(fasting glucose mg/dL)). HOMA2 IR takes account of variations in hepatic and peripheral glucose resistance, increases in the insulin secretion curve for plasma glucose concentrations above 10 mmol/L (180 mg/dL) and the contribution of circulating proinsulin[21]. HOMA2 is also calibrated to give %B and %S values of 100% in normal young adults when using currently available assays for insulin, specific insulin or C-peptide[21].

### 2.6 Intervention adherence

Secoiridoids, such as olecanthal and oleacein, the two main polyphenols in the selected oils for this study, are known to be partly hydrolyzed in the stomach, resulting in a significant increase in their derivates, i.e., free tyrosol and hydroxytyrosol, respectively. Hence, we decided to measure the levels of one of these derivatives in the plasma of patients in order to assess their adherence to the nutritional intervention. We selected hydroxytyrosol because their quantification in our hands is more accurate and precise than that of tyrosol. To determine the total content of hydroxytyrosol in the plasma samples, an acidic hydrolysis was carried out. The phenolic extraction was achieved based in previus reports[22,23] by solid phase extraction (SPE) in microelution plates, Oasis HLB (Waters, Milford, USA). Samples were eluted with 125 µl methanol, filtered through 0.45 µm filters and injected into the UHPLC-MS. The samples were separated in a Dionex Ultimate 3000RS UHPLC (Thermo Fisher Scientific, Waltham, MA, USA), equipped with a quaternary pump, autosampler and a photodiode array detection (DAD) system. Chromatographic separation was performed on a Mediterranea SEA18 column (200 x 4 mm i.d., 3µm particle size, Teknokroma, Barcelona, Spain) at 30 °C. Finally, to determine the structures of the compounds a micrOTOF-Q II^TM^ High Resolution Time-of-Flight mass spectrometer (UHR-qTOF) with Q-q-TOF geometry (Bruker Daltonics, Bremen, Germany) and equipped with an electrospray ionization (ESI) source operating in negative ion mode was used. Instrument control and data evaluation were performed with Bruker Daltonics HyStar 3.2 and Bruker Daltonics DataAnalysis 4.2., respectively.

### 2.7 Inflammatory cytokines and oxidative stress measurement

Inflammatory cytokines (adiponectin, leptin, TNF-α, IFN-γ, CXCL1, IL-10, IL-12p40, IL-13, IL-1RA, IL-1β, IL-4, IL-6) were measured with ProcartaPlex multiplex Immunoassay (Thermo Fisher Scientific, Waltham, MA, USA) following manufacturer’s instructions. Total Antioxidant Status (TAS) and Glutathione Reductase (GRD) were determined using a colorimetric and ultraviolet assay kit, respectively (Randox Laboratories Ltd, Crumlin, UK). Determination of plasma sulfhydryl groups (total thiols) was done by using Ellman’s reagent 5,5′-dithiobis (2-nitrobenzoate)-DTNB adapted to a ICubio Autoanalyzer and -SH concentration was calculated by using a standard curve of glutathione. Aqueous and lipid hydroperoxides were determined using the peroxidetect kit (Sigma-Aldrich Co. LLC) following manufacturer’s instructions.

### 2.8 Ethics

The protocol was approved on Nov 29, 2017 by the Research Ethic Committee of Málaga (Spain). All participants signed the informed consent upon explanation of all the objectives and methodology of the trial. The APRIL study (Clinical Trial Registration: ISRCTN17232860 https://doi.org/10.1186/ISRCTN17232860) was conducted according to the recommendations of the Helsinki Declaration and Good Clinical Practice Guidelines of the International Council for Harmonization (CPMP/ICH/135/95) and the current Spanish directives (RD 1090/2015).

### 2.9 Sample size calculation and statistical analysis

Based on the reported differences in the inflammatory marker C-reactive protein (CRP) between healthy and obese people we calculated a sample size of 130 individuals, 65 in each group that would enable the study to have 80% power to detect differences between the two groups in this clinical marker related to inflammation, with a two-sided significance level of 5% and a 15% drop-out rate [24]. Frequency distributions were used to describe qualitative outcome measures, while quantitative measures were described using means and standard deviations. The parametricity of the variables was examined and logarithmic transformation of the variables was performed if required. Differences in the baseline characteristics among sequences of administration were assessed by an ANOVA test. Differences among treatments were assessed by an ANCOVA test adjusted by age, sex, and sequence of treatments administration. A p value of <0.05 was considered significant. All data were analyzed using SPSS V.26.0 for Windows (SPSS Inc., Chicago, Illinois, USA).

## 3. Results

### 3.1 Recruitment, baseline characteristics of participants and compliance

More than 400 subjects were selected as potential participants and contacted for a recruitment visit. 264 patients attended this recruitment visit and 91 of them met the criteria and agreed to participate in the study, signing the informed consent (Figure 1). The recruitment took place between March 2018 and February 2019. No dropouts were recorded during the trial. No differences were observed in baseline characteristics of the participants by sequence of intervention (Table 2) with the exception of a lower physical activity in those following sequence 2 (EVOO followed by OO).

**Table 2.** Baseline characteristics of participants by sequence of administration

|  | Sequence 1<br>(n = 46) | Sequence 2<br>(n=45) | P |
| --- | --- | --- | --- |
| Age. y | 56.1 ± 6.12 | 54.9 ± 5.36 | 0.315 |
| Female. % | 63.0 | 64.4 | 0.532 |
| Civil status |  |  | 0.236 |
| Single | 17.4 | 4.4 |  |
| Married/Partner | 69.6 | 84.4 |  |
| Widow | 4.3 | 4.4 |  |
| Divorced/Separated | 8.7 | 6,7 |  |
| Smoking habits. % |  |  | 0.618 |
| Never | 36.9 | 46.7 |  |
| Smoker | 26.1 | 20.0 |  |
| Exsmoker | 36.9 | 33.3 |  |
| Systolic blood pressure, *mm Hg* | 132 ± 16.6 | 130 ± 15.2 | 0.340 |
| Diastolic blood pressure, *mm Hg* | 77 ± 8.79 | 75 ± 10.9 | 0.357 |
| Pulse pressure, *mm Hg* | 56 ± 12.5 | 53 ± 11.4 | 0.081 |
| Weight, *kg* | 93 ± 14.4 | 89 ± 12.1 | 0.210 |
| Body mass index, *kg/m<sup>2</sup>* | 34.8 ± 4.27 | 33.7 ± 3.61 | 0.179 |
| Waist circumference, *cm* | 109 ± 10.6 | 107 ± 10.0 | 0.435 |
| Waist/Hip, *cm* | 0.96 ± 0.09 | 0.95 ± 0.08 | 0.822 |
| Waist/height | 0.67 ± 0.06 | 0.66 ± 0.06 | 0.477 |
| Total Physical Activity<sup>#</sup> | 536 (198-1386) | 313 (1-1254) | 0.048* |
| Mediterranean Diet Score | 8.3 ± 1.93 | 8.6 ± 2.28 | 0.537 |

Sequence 1: OO followed by EVOO 2; Sequence 2: EVOO followed by OO. Data expressed as mean ± standard deviation, or percentages. <sup>#</sup> median (25<sup>th</sup>-75<sup>th</sup> percentiles). P for ANOVA, Mann Whitney, or $\chi^2$ tests.

Adherence to treatment was assessed by measuring changes in plasma levels of hydroxityrosol, as previously mentioned. Hydroxytyrosol levels increased after EVOO intervention, suggesting compliance with EVOO treatment, but not after OO intervention, probably reflecting no enough differences in the content of this polyphenol between OO and the oil used before the study (Figure 2). This latter finding was expected as most people in Spain consume OO as the primary source of lipids in diet. No inter-treatment differences were observed.

**Figure 2.**
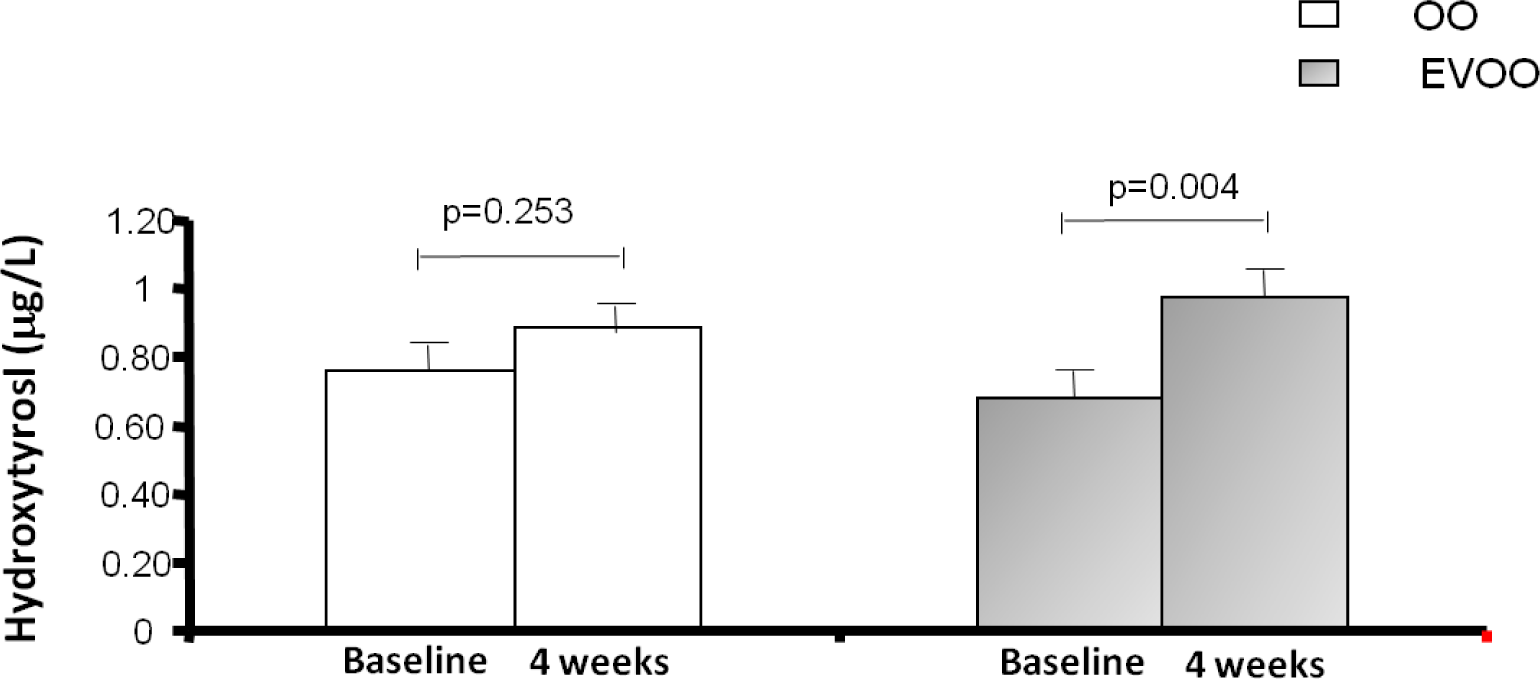
Plasma hydroxytyrosol (µg/L) before and after treatments. Data expressed as mean (SE). Student’s t test for related samples.

### 3.2 Food intake and Mediterranean Diet Score during the trial

Supplementary Table 1 shows the changes in groups of food consumption during the trial. No inter-treatment differences were observed. After OO treatment there was a decrease (p=0.043) in white meat consumption and a decrease in alcohol (p=0.026) were observed after EVOO treatment. After both treatments decreases in solid fats (p<0.05), vegetables in stew (p<0.005), and an increase in olive oil consumption (p< 0.005) were observed. Neither intra- nor inter-treatments differences were observed in the Mediterranean Diet Score after treatments (Supplementary Table 2).

### 3.3 Physical activity, blood and pulse pressure and anthropometric and adiposity parameters

No changes in physical activity after 4 weeks of treatment were observed either intra- or inter-treatments (Supplementary Table 3). No changes in blood and pulse pressure were found either (Supplementary Table 4). However, there was a significant decrease in weight and in the BMI after the EVOO treatment (p<0.05), although differences between treatments did not reach significance. Neither intra- nor inter-treatments differences were observed in other anthropometric and adiposity markers (Table 3).

**Table 3.**
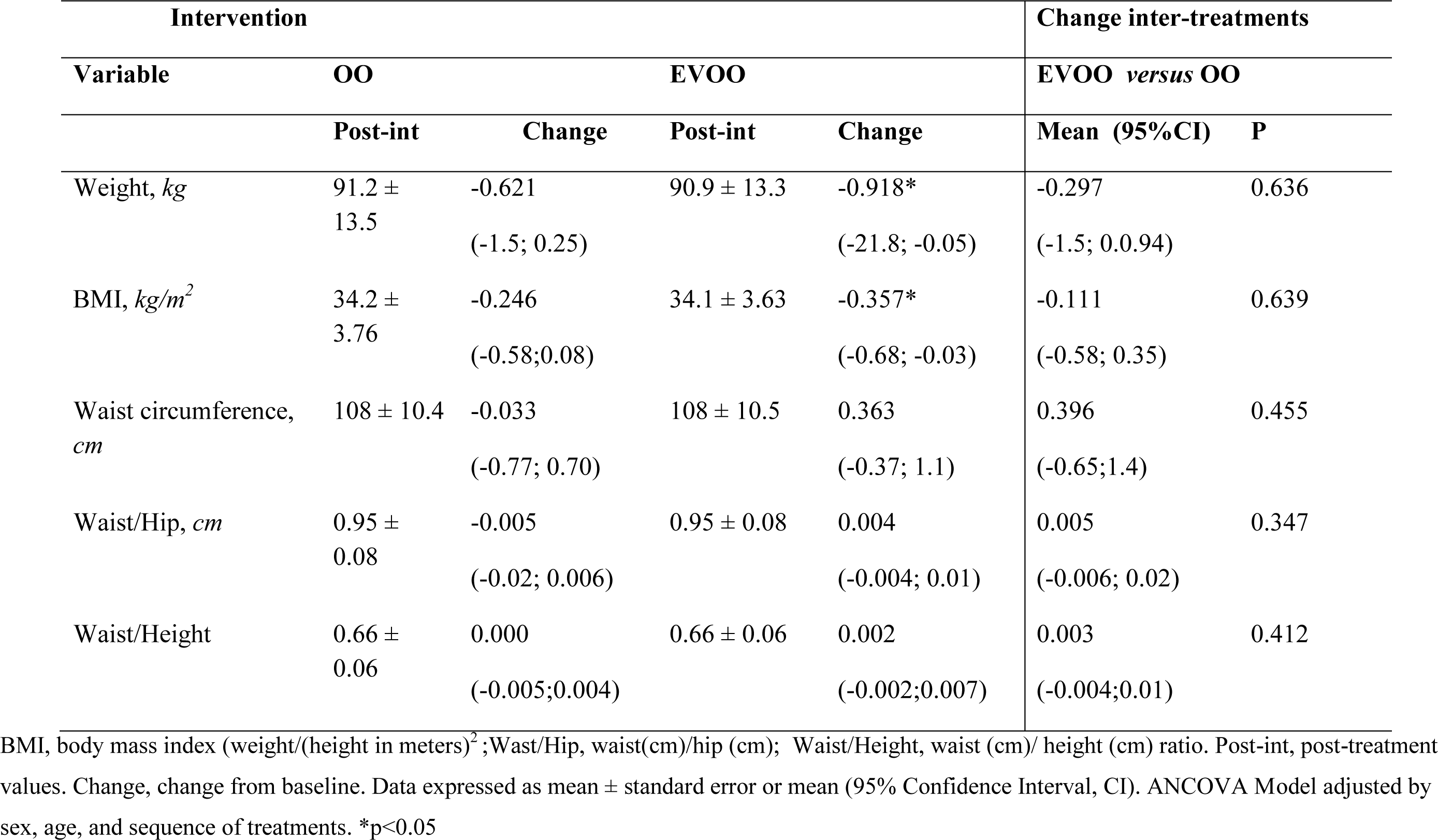
Changes in anthropometric and adiposity measures at 4weeks of intervention

| Intervention |  |  |  |  | Change inter-treatments |  |
| --- | --- | --- | --- | --- | --- | --- |
| Variable | OO |  | EVOO |  | EVOO *versus* OO |  |
|  | Post-int | Change | Post-int | Change | Mean (95%CI) | P |
| Weight, *kg* | 91.2 ± 13.5 | -0.621<br>(-1.5; 0.25) | 90.9 ± 13.3 | -0.918*<br>(-21.8; -0.05) | -0.297<br>(-1.5; 0.0.94) | 0.636 |
| BMI, *kg/m<sup>2</sup>* | 34.2 ± 3.76 | -0.246<br>(-0.58;0.08) | 34.1 ± 3.63 | -0.357*<br>(-0.68; -0.03) | -0.111<br>(-0.58; 0.35) | 0.639 |
| Waist circumference, *cm* | 108 ± 10.4 | -0.033<br>(-0.77; 0.70) | 108 ± 10.5 | 0.363<br>(-0.37; 1.1) | 0.396<br>(-0.65;1.4) | 0.455 |
| Waist/Hip, *cm* | 0.95 ± 0.08 | -0.005<br>(-0.02; 0.006) | 0.95 ± 0.08 | 0.004<br>(-0.004; 0.01) | 0.005<br>(-0.006; 0.02) | 0.347 |
| Waist/Height | 0.66 ± 0.06 | 0.000<br>(-0.005;0.004) | 0.66 ± 0.06 | 0.002<br>(-0.002;0.007) | 0.003<br>(-0.004;0.01) | 0.412 |

BMI, body mass index (weight/(height in meters)<sup>2</sup>; Wast/Hip, waist(cm)/hip (cm); Waist/Height, waist (cm)/ height (cm) ratio. Post-int, post-treatment values. Change, change from baseline. Data expressed as mean ± standard error or mean (95% Confidence Interval, CI). ANCOVA Model adjusted by sex, age, and sequence of treatments. \*p<0.05

### 3.4 Lipid variables and glucose homeostasis parameters

No changes in lipid variables were found (Supplementary Table 5). Neither intra- nor inter-treatments differences were observed in triglycerides, total cholesterol, HDL cholesterol and LDL cholesterol as well as in the atherogenic ratios total cholesterol/HDL, LDL/HDL and triglycerides/HDL. Table 4 shows the changes in glucose homeostasis parameters. Interestingly, glucose levels were significantly decreased after EVOO treatment but not after OO treatment, although inter-treatments differences did not reach statistical significance. Neither intra- nor inter-treatments differences were observed in insulin, HOMA-IR, QUICKI, HOMA2 and HbA1c.

**Table 4.** Changes in glucose homeostasis parameters at 4 weeks of interventions

| Intervention |  |  |  |  | Change inter-treatments |  |
| --- | --- | --- | --- | --- | --- | --- |
| Variable | OO |  | EVOO |  | EVOO *versus* OO |  |
|  | Post-int | Change | Post-int | Change | Mean (95%CI) | P |
| Glucose, *mg/dL* | 93 ± 16.8 | -2.05<br>(-5.7; 1.6) | 93± 20.8 | -3.69*<br>(-7.3; -0.05) | -1.63<br>(-6.8; 3.5) | 0.530 |
| Insulin, *mUI/mL* | 14.8 ± 10.2 | -0.618<br>(-3.2;2.0) | 15.1 ± 9.95 | -0.213<br>(-2.8; 3.4) | 0.404<br>(-3.3; 4.1) | 0.828 |
| HOMA-IR | 3.46± 2.52 | -0.159<br>(-0.86; 0.54) | 3.95 ± 4.30 | -0.168<br>(-0.87; 0.54) | -0.009<br>(-1.0;0.99) | 0.986 |
| QUICKI | 0.33 ± 0.03 | 0.001<br>(-0.005;0.006) | 0.32 ± 0.03 | -0.001<br>(-0.006;0.005) | -0.001<br>(-0.009;0.007) | 0.749 |
| HbA1c, % | 5.81± 0.33 | -0.005<br>(-0.04;0.03) | 5.80± 0.35 | -0.020<br>(-0.06;0.02) | -0.015<br>(-0.07;0.04) | 0.558 |
| HOMA2%B (log) | 2.10 ± 0.23 | 0.010<br>(-0.04; 0.06) | 2.15 ± 0.24 | 0.043<br>(-0.005;0.09) | 0.033<br>(-0.03;0.10) | 0.344 |

**Table 4 (cont)**
|  |  |  |  |  |  |  |
| --- | --- | --- | --- | --- | --- | --- |
| HOMA2% S (log) | 1.80 ± 0.22 | 0.000<br>(-0.04;0.04) | 1.74 ± 0.24 | -0.039<br>(-0.08;0.04) | -0.039<br>(-0.10;0.02) | 0.197 |
| HOMA2_IR (log) | 1.79 ± 0.91 | -0.094<br>(-0.39;0.20) | 1.91 ± 0.97 | 0.267<br>(-0.03;0.57) | 0.360<br>(-0.06;0.78) | 0.093 |

HOMA-IR, homeostatic model assessment for insulin resistance, calculated as: (Fasting Insulin x Fasting Glucose)/405; QUICKI, quantitative insulin sensitivity index, calculated as $1 / ((\log (\text{fasting insulin } \mu\text{U/mL}) + \log(\text{fasting glucose mg/dL}))$ ; HbA1c, Glycated hemoglobin; HOMA2: calculator: HOMA2%B, homeostatic model assessment for beta-cell function; HOMA2% S homeostatic model assessment for insulin sensitivity; HOMA 2\_IR. Data expressed as mean ± standard deviation or mean (95% Confidence Interval, CI). ANCOVA Model adjusted by sex, age, and sequence of treatments administration. \*p<0.05

### 3.5 C reactive protein, thyrotropin, 25-OH-vitamin D and renal function parameters

No changes in C reactive protein (both data of the high sensibility test or not), thyrotropin, and 25-OH-vitamin D at 4 weeks of interventions were detected (Supplementary Table 6). There were no inter-treatments differences in these parameters either. Regarding renal function, no differences were observed either intra- or inter-treatments in urea, creatinine, or glomerular filtration rate. Uric acid decreased after both treatments reaching significance after the OO treatment (Supplementary Table 7). No inter-treatments differences were observed.

### 3.6 Inflammatory cytokines

Table 5 shows the changes in inflammatory cytokines after 4 weeks of treatments. EVOO treatment significantly decreased the interferon gamma, both intra-treatment and when changes where compared with those obtained after OO treatment (p=0.041). Both treatments increased CXCL-1. The increase after EVOO treatment, however, was lower, with a borderline significance (p=0.080) when comparing with changes after OO treatment. IL-12p40 and IL-1RA increased after both treatments without inter-treatment differences. Neither intra- nor inter-treatments differences were observed in other cytokines.

**Table 5.** Changes in inflammatory cytokines at 4 weeks of interventions

| Intervention |  |  |  |  | Change inter-treatments |  |
| --- | --- | --- | --- | --- | --- | --- |
| Variable | OO |  | EVOO |  | EVOO *versus* OO |  |
|  | Post-int | Change | Post-int | Change | Mean (95%CI) | P |
| Adiponectin (log), $\mu\text{g/mL}$ | $0.55 \pm 0.25$ | -0.027<br>(-0.07; 0.02) | $0.53 \pm 0.35$ | -0.040<br>(-0.09; 0.009) | -0.013<br>(-0.08; 0.06) | 0.704 |
| Leptin (log), $\text{pg/mL}$ | $3.75 \pm 0.31$ | 0.000<br>(-0.02;0.02) | $3.76 \pm 0.32$ | -0.006<br>(-0.03;0.02) | -0.006<br>(-0.04;0.03) | 0.733 |
| TNF-α, $\text{pg/mL}$ | $10.5 \pm 3.03$ | 0.036<br>(-0.20;0.27) | $10.3 \pm 2.78$ | 0.008<br>(-0.29;0.24) | -0.029<br>(-0.36; 0.30) | 0.866 |
| IFN-γ (log), $\text{pg/mL}$ | $1.52 \pm 0.06$ | 0.002<br>(-0.004; 0.008) | $1.52 \pm 0.07$ | -0.007*<br>(-0.01; -0.001) | -0.009<br>(-0.02; -0.000) | 0.041* |
| CXCL1 (log), $\text{pg/mL}$, (N=77)<br>*Samples with non-detectable values were omitted* | $0.80 \pm 0.23$ | 0.336*<br>(0.27;0.40) | $0.66 \pm 0.35$ | 0.251*<br>(0.18;0.32) | -0.085<br>(-0.18;0.01) | 0.080 |

**Table 5 (cont)**
|  |  |  |  |  |  |  |
| --- | --- | --- | --- | --- | --- | --- |
| IL-10, pg/mL | 6.84 ± 1.49 | 0.089<br>(-0.08;0.25) | 6.71 ± 1.43 | -0.072<br>(-0.24; 0.09) | -0.160<br>(-0.39; 0.07) | 0.179 |
| IL-12p40 (log), pg/mL (N=64)<br>*Samples with non-detectable values were omitted* | 0.42 ± 0.31 | 0.122*<br>(0.07;0.17) | 0.44 ± 0.31 | 0.164*<br>(0.11;0.22) | 0.037<br>(-0.03; 0.12) | 0.255 |
| IL-13, pg/mL | 4.52 ± 0.51 | 0.000<br>(-0.06;0.06) | 4.44 ± 0.43 | -0.036<br>(-0.10;0.02) | -0.039<br>(-0.13; 0.05) | 0.396 |
| IL-1RA (log), pg/mL (N=77)<br>*Samples with non-detectable values were omitted* |  | 0.339*<br>(0.26;0.42) |  | 0.265*<br>(0.18;0.35) | -0.073<br>(-0.19; 0.42) | 0.210 |
| IL-1β (log), pg/mL | 0.76 ± 0.13 | 0.000<br>(-0.01;0.01) | 0.76 ± 0.14 | -0.007<br>(-0.02;0.06) | -0.007<br>(-0.02; 0.08) | 0.485 |
| IL-4, pg/mL | 23.3 ± 2.13 | 0.098<br>(-0.14;0.34) | 23.3 ± 2.18 | 0.089<br>(-0.15;0.33) | -0.010<br>(-0.35; 0.33) | 0.955 |
| IL-6, pg/mL | 42.8 ± 14.2 | 0.192<br>(-0.94;1.-3) | 42.1 ± 13.1 | 0.096<br>(-1.3;1.2) | -0.095<br>(-1.7; 1.5) | 0.906 |

Data expressed as mean ± standard deviation or mean (95% Confidence Interval, CI) (n=91). Logarithmic transformation of the data for their normalization if required. TNF-α, tumor necrosis factor-alpha; IFN-γ, interferon gamma; CXCL1, chemokine (C-X-C motif) ligand 1; IL, interleukin; IL-1RA, interleukin-1 receptor antagonist. ANCOVA Model adjusted by sex, age, and sequence of treatments administration. \*p<0.05

### 3.7 Oxidative stress parameters

Table 6 shows the changes in oxidative stress parameters. EVOO treatment significantly increased the total antioxidant status, both intra-treatment and when changes where compared with those obtained after OO treatment (p=0.043). Total Thiols decreased after OO treatment. When comparing inter-treatment changes, total Thiols increased after EVOO versus OO treatment with a borderline significance (p=0.091). EVOO treatment decreased lipid peroxides, both the hydroxy and the organic ones, the decreases being significant in both cases versus changes after OO treatment (p=0.011 and p=0.008, respectively). Both treatments decreased Glutathione reductase without inter-treatment changes.

**Table 6.** Changes in oxidative stress parameters at 4 weeks of interventions

| Intervention |  |  |  |  | Change inter-treatments |  |
| --- | --- | --- | --- | --- | --- | --- |
| Variable | OO |  | EVOO |  | EVOO *versus* OO |  |
|  | Post-int | Change | Post-int | Change | Mean (95%CI) | P |
| Total Antioxidant Status, *mmol/L* | 1.48 ± 0.31 | -0.019<br>(-0.06; 0.03) | 1.51 ± 0.32 | 0.048*<br>(0.002; 0.9) | 0.067<br>(0.002; 0.131) | 0.043* |
| Glutathione reductase, *U/L* | 45.4 ± 12.2 | -3.16*<br>(-5.5;-0.78) | 45.2 ± 13.3 | -3.46*<br>(-5.8;-1.1) | -0.300<br>(-3.6; 3.1) | 0.860 |
| Total Thiols<sup>a</sup>, μM | 102 ± 22 | -6.94*<br>(-12; -2.2) | 106 ± 26 | -1.25<br>(-5.9; 3.4) | 5.69<br>(-0.92; 12) | 0.091 |
| Hydro-LOOH<sup>b</sup>, μM | 0.52 ± 0.22 | 0.028<br>(-0.006;0.06) | 0.48 ± 0.26 | -0.035*<br>(-0.07;-0.001) | -0.063<br>(-0.11;-0.01) | 0.011* |
| Organic -LOOH<sup>c</sup>, μM | 9.29 ± 4.45 | 0.364<br>(-0.07;0.69) | 8.49 ± 5.42 | -0.462*<br>(-0.88;-0.04) | -0.826<br>(-1.4;-0.22) | 0.008** |

Data expressed as mean ± standard deviation or mean (95% Confidence Interval, CI). <sup>a</sup> Albumin adjusted; <sup>b</sup> Lipid hydroperoxides, adjusted by triglycerides; <sup>c</sup> Organic lipoperoxides, adjusted by triglycerides. ANCOVA Model adjusted by sex, age, and sequence of treatments administration. \*p<0.05, \*\*p<0.01

## 4. Discussion

Overall, the main findings arising from this study are: 1) A nutritional intervention with an EVOO rich in oleacein and oleocanthal induces a systemic potent antioxidant effect in obese people with prediabetes when compared to a OO with a similar content of simple polyphenols; 2) This antioxidant effect is accompanied by a differential mild anti-inflammatory effect and 3) a significant decrease in body weight, BMI and glycemia, all of them well-recognized risk factors for the development of T2D, metabolic syndrome and cardiovascular diseases.

In order to specifically assess the health effects of these olive oils, we intended to be as less interventional as possible. For this purpose, no fixed amounts of olive oil ingestion were requested and no advices on physical activity or changes in any other habit were proposed. In fact, no changes were detected in the MedDiet score and only minor changes were detected in food intake, some of them related to the requested change in the source of lipids. Thus, the amount of solid fats decreased in both groups while that of olive oil increased. We also detected a decrease in vegetables in stews in both groups, and a decrease in white meat in OO and in alcohol in EVOO group. The reasons for these changes in vegetables, white meat and alcohol ingestion, apparently unrelated to changing the lipid source, are unknown. No changes in physical activity were detected as per the IPAQ questionnaire results. Therefore, in terms of intervention we believe that our results represent the biological response to a change in the main lipid source with minor, if any, interference of changes in diet or physical activity.

Similarly, the good adherence of patients to this intervention is highlighted by the facts that there were no withdrawals (all the enrolled patients finished the trial) and the levels of hidroxytyrosol in plasma (a simple polyphenol but also a metabolite of oleacein) were higher after the EVOO treatment. As mentioned before in the result section, no significant changes were expected in hidroxytyrosol levels after OO treatment as these people, living in a Mediterranean country, already use olive oil as the main source of lipid in diets.

The main differential effect elicited by EVOO when compared to OO was a clear improvement in oxidative stress at the systemic level. Specifically, EVOO increased the anti-oxidant status thus increasing the capacity to respond to an oxidative challenge, decreased the activity of glutathione reductase what suggest a lower demand of reduced glutathione, maintained the levels of total thiols, the main antioxidants in the body, and decreased lipid peroxidation, as assessed by measuring lipid peroxides both in the soluble and the organic fraction. Only a decrease in glutathione reductase activity and a decrease in total thiols were detected after OO treatment. The decrease in total thiols could be related to a higher use of this antioxidant in people treated with an olive oil low in secoiridiods, and the paradoxical decrease in glutathione reductase activity could be reflecting changes in the regulation of this enzyme by inflammatory and hepatic factors[25]. Therefore, one-month intervention with EVOO promoted a healthier oxidative profile than OO in obese people with prediabetes. The bioactive compounds involved in these beneficial effects of EVOO could be the complex polyphenols, especially oleocanthal and oleacein which have been demonstrated to elicit some anti-oxidant effects in *in vitro* and *in vivo* assays[26–30] and constitute 90% of total polyphenol content in this EVOO. However, a contribution of other minor complex polyphenols such as oleuropein, ligstroside as well as lignans and flavonoids, at least partially, cannot be ruled out. Interestingly, oxidative stress is increasingly becoming recognized as a key process in diabetes pathophysiology, specifically in beta cell failure, the driver of diabetes onset[31,32]. Hence, olive oils rich in oleocanthal and oleacein, able to counteract oxidative stress, might mediate prevention or delay in the progression from prediabetes to diabetes, though this aspect need further clinical validation.

Another important process involved in obesity pathophysiology and related complications, including diabetes, is inflammation. In our study, where the primary outcome was the inflammatory status, we found an overall mild effect on inflammation, with the nutritional interventions being able to modulate some inflammatory markers. Specifically, CXCL1, IL-12p40 and IL-1RA were increased after both treatments. CXCL1 and IL-12p40 are immunomodulatory molecules which have been related to T2D and other inflammatory conditions [33,34], acting as chemoattractant for several immune cells, especially neutrophils and macrophages, respectively. In addition, IL-12p40 provides a negative feedback loop by competitively binding the pro-inflammatory IL-12 receptor [34]. On the other side, IL-1RA is a natural anti-inflammatory factor and mediator in glucose homeostasis disturbances that blocks IL-1α and IL-1β signalling[35]. While the increase in CXCL1 after olive oil treatments is puzzling given the current lack of studies on the role of this small peptide in obesity and diabetes pathogenesis[33], those of IL-12p40 and IL-1RA fits well with an anti-inflammatory effect mediated by blockage of pro-inflammatory signalling through IL-12 and IL-1 receptors, respectively[34,35]. Interestingly, IL-12p40 has also been found to suppress IFN-γ secretion [36], a cytokine produced during chronic inflammation and also related to the events initiating obesity-induced adipose tissue inflammation and insulin resistance[37]. IFN-γ is a master regulator of the immune response and control, among other processes, the production of pro-inflammatory cytokines. Of note, EVOO specifically decreased the levels of IFN-γ both intra- and inter-group, suggesting a higher capacity to modulate systemic inflammation when compared to OO. Oleocanthal and oleacein could be mediating this latter effect because their anti-inflammatory actions are well-documented[8,9]. In fact, an interventional study for two months with high oleocanthal EVOO in people with metabolic syndrome and hepatic steatosis found improvements in the inflammatory cytokines IL-6, IL17A, TNF-α, IL-1β and IL-10[14]. We did not detect relevant changes in these cytokines but it may be the case that one month of treatment is insufficient to induce robust changes in some cytokines. Indeed, the lower than expected number of patients recruited (sample size estimated using the hsCRP variable), could explain the lack of detection of some changes in inflammatory markers. Overall, our results suggest a mild anti-inflammatory effect of nutritional interventions, being higher in that performed with EVOO.

Interestingly, EVOO but not OO was able to significantly decrease body weight (close to one kilogram after one month of treatment) and BMI, a finding that was paralleled by an improvement in fasting glucose. The improvement in glucose homeostasis was not accompanied by changes in insulin resistance as assessed by several surrogated markers. No changes were detected in blood pressure and lipid parameters either. Taken together, these findings suggest that EVOO was able to induce some clinical improvements in glucose handling, probably related with body weight decrease and amelioration of the inflammatory and oxidative status. These findings also agree with those reported in an intervention with high oleocanthal EVOO on patients with metabolic syndrome and hepatic steatosis, where the intervention reduced body weight, BMI and waist circunference[14].

The main strengths in this study are the crossover nature of the trial, thus allowing all the subjects to be analyzed before and after both treatments, the specificity of the intervention, avoiding interferences of changes in diet and physical activity, and the good adherence to the treatment.

This study has also several limitations that have to be taken into account. First, the number of patients recruited was lower than expected, thus eventually limiting the detection of changes in some variables. Second, due to budget constraints, the duration of each intervention was short (one month) what might have precluded the detection of more striking differences. Finally, the clinical relevance of these findings is limited as, due to the design and short duration of the trial, we could not estimate the incidence of diabetes or other metabolic complications in these patients.

In conclusion, this study shows that a nutritional intervention with high oleocanthal and oleacein EVOO induces a healthier profile than one with OO in people with obesity and prediabetes, promoting body weight loss, improving glucose homeostasis and ameliorating the inflammatory and oxidative status. These findings are relevant in terms of designing MedDiet-based nutritional interventions for people at risk of developing diabetes and related comorbidities.

## Data Availability

All data produced in the present study are available upon reasonable request to the authors

## Acknowledgements

The authors want to thank Sociedad Andaluza de Oleocantal and the International Oleocanthal Society, based in Benalmádena (Málaga), and specially Dr. José Antonio Amérigo and Clara Villanueva for their continuous support and contact with mills and olive oil producers. The authors are also indebted with Manuel Heredia Halcón from Cortijo Suerte Alta S.L. (Albendín, Baena, Córdoba, Spain) and Justino Damián Corchero from Jacoliva S.L. (Pozuelo de Zarzón, Cáceres, Spain) for the free and altruistic supply of EVOO. OO was purchased from Aceites Albert S.A. (Valencia, Spain). Dr. Gustavo Fernández Usero, director of the primary care center at Alhaurin El Grande, Málaga, for his kindness and help in recruiting patients. Vanesa Espinosa-Jiménez, for technical support. NUPROAS HB, for outstanding statistical advice and data processing and analysis. A special thanks to all the patients who voluntarily and altruistically participated in this study.

## Author contribution

FJBS, SYRZ, GOF and GRM contributed to the study design. FJBS obtained funding. IRG, MG, SV, MJTG, FMC, MCC and GOF recruited patients. RB and IRG collected data and samples. RB, MGF, SYRZ, EL, EMM, ROF, REB, ASG and AGB processed samples. FJBS, MIC, GRM, IRG, JCF and AGB contributed to data analysis and interpretation. FJBS, SYRZ and IRG wrote the manuscript. All the authors reviewed and commented on the manuscript. FJBS is the guarantors of this work and takes responsibility for the integrity of the data and the accuracy of the data analysis.

## Data accessibility statement

The datasets generated during the trial are available from the corresponding author upon reasonable request.

---

**Supplementary Table 1.**
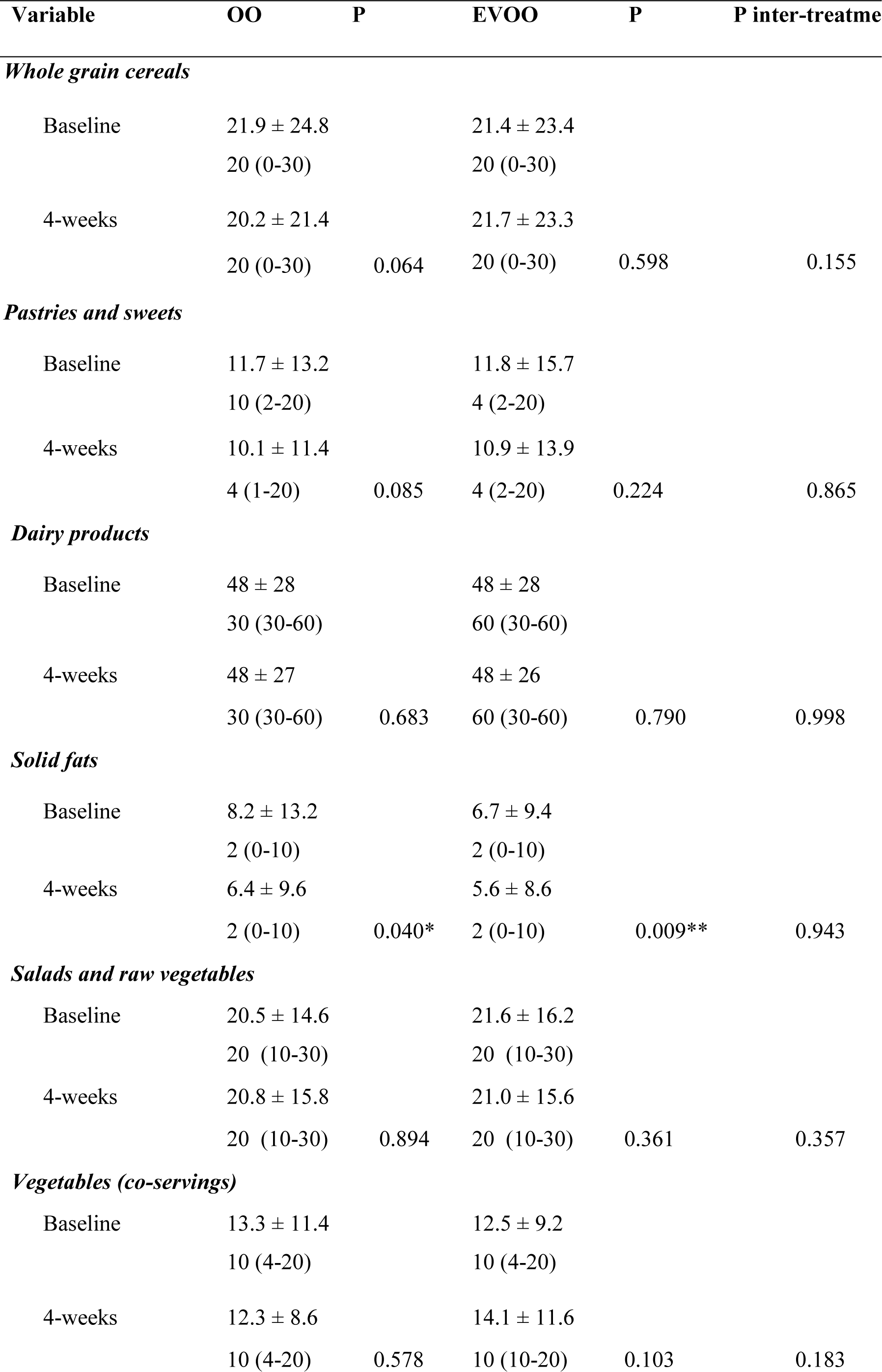
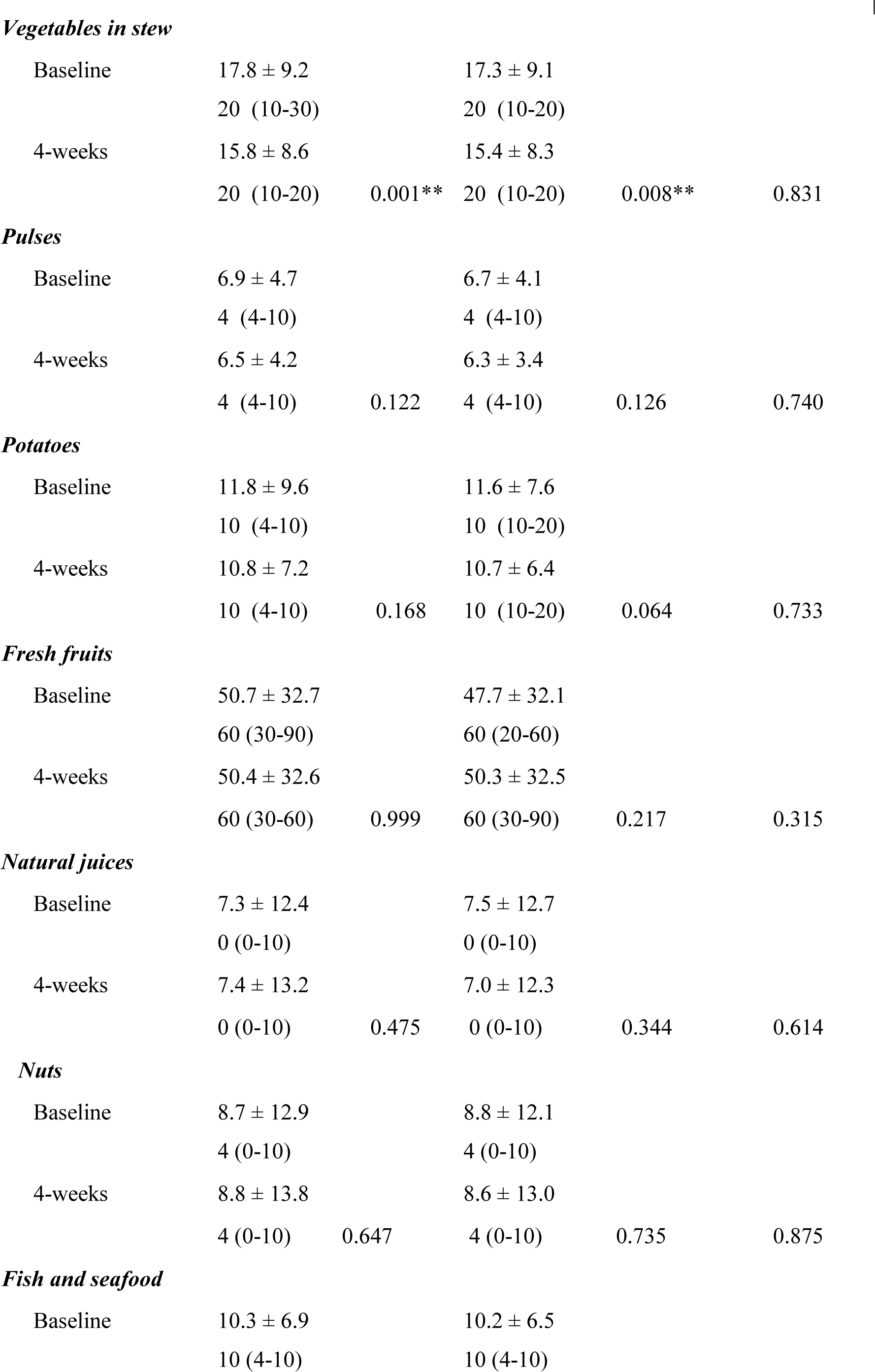
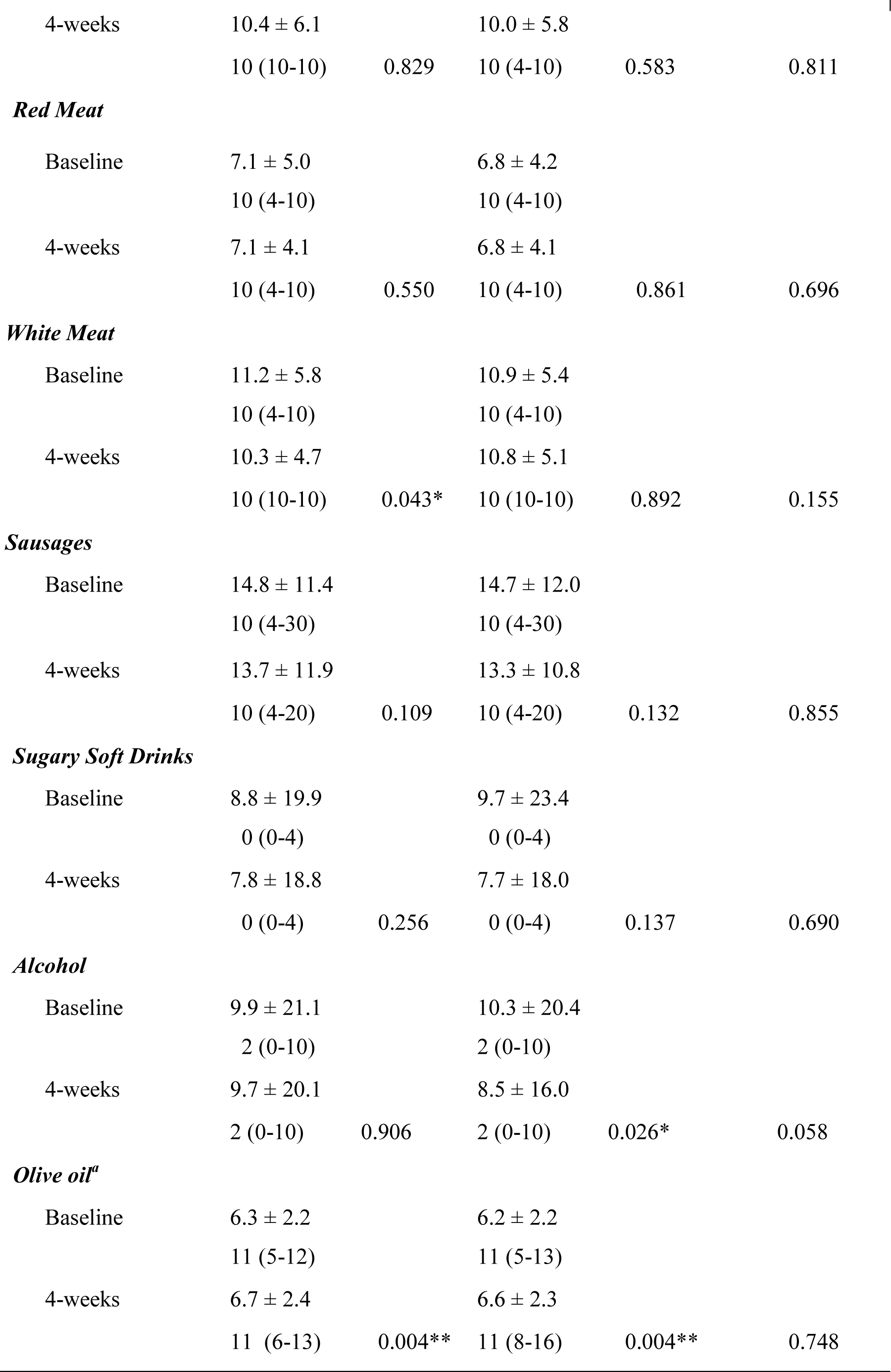
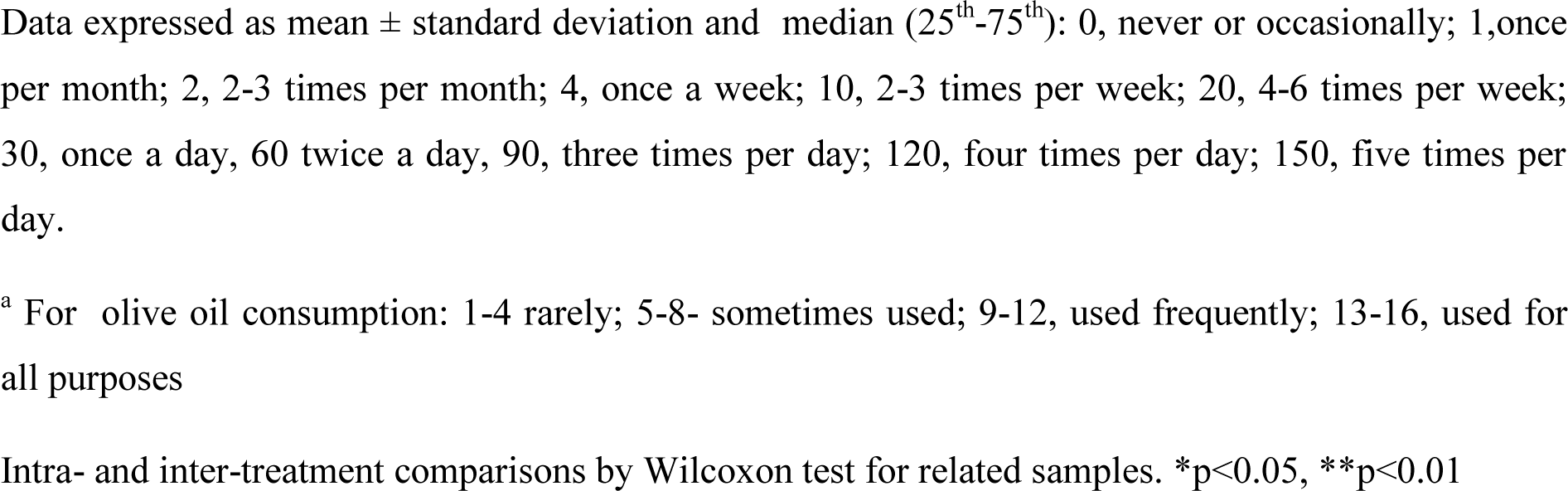
Changes in Food Groups after 4 weeks of interventions

**Supplementary Table 2.** Changes in Mediterranean Diet (MedDiet) Score after 4 weeks of treatment

|  | Intervention |  |  |  | Change inter-treatments |  |
| --- | --- | --- | --- | --- | --- | --- |
| Variable | OO |  | EVOO |  | EVOO *versus* OO |  |
|  | Post | Change | Post | Change | Mean<br>(95%CI) | P |
| MedDiet Score | 8.6 ± 2.101 | 0.077<br>(-0.20;0.35) | 8.2 ± 2.18 | 0.022<br>(-0.25;0.30) | -0.055<br>(-0.44;0.33) | 0.780 |

Post, post-intervention values. Change, change from baseline. Data expressed as mean ± standard deviation or mean (95% Confidence Interval, CI). ANCOVA Model adjusted by sex, age, and sequence of treatments administration. No differences were observed.

**Supplementary Table 3.** Changes in Physical Activity (MET-minutes/week) after 4weeks of interventions

| Variable | OO | P | EVOO | P | P for difference |
| --- | --- | --- | --- | --- | --- |
| Walking |  |  |  |  |  |
| Baseline | 330 (0-792) |  | 247 (33-693) |  |  |
| 4-weeks | 330 (132-792) | 0.117 | 330 (99-594) | 0.785 | 0.387 |
| Moderate |  |  |  |  |  |
| Baseline | 0 (0-120) |  | 0 (0-80) |  |  |
| 4-weeks | 0 (0-0) | 0.410 | 0 (0-240) | 0.816 | 0.791 |
| Vigorous |  |  |  |  |  |
| Baseline | 0 (0-0) |  | 0 (0-0) |  |  |
| 4-weeks | 0 (0-0) | 0.999 | 0 (0-0) | 0.715 | 0.944 |
| Total physical activity |  |  |  |  |  |
| Baseline | 445 (49-1386) |  | 396 (39-1200) |  |  |
| 4-week | 445 (198-1158) | 0.757 | 594 (148-1272) | 0.624 | 0.973 |

Data expressed as (median, 25<sup>th</sup>-75<sup>th</sup> percentile). Moderate, moderate physical activity: those physical activities that require moderate physical effort such as carrying light things, riding a bicycle at a regular pace, or playing doubles in tennis Vigorous, vigorous physical activity: those physical activities that require a strong physical effort such as lifting heavy objects, digging, aerobics, or cycling fast.

**Supplementary Table 4.** Changes in blood and pulse pressure at 4weeks of interventions

| Intervention |  |  |  |  | Change inter-treatments |  |
| --- | --- | --- | --- | --- | --- | --- |
| Variable | OO |  | EVOO |  | EVOO *versus* OO |  |
|  | Post-int | Change | Post-int | Change | Mean (95%CI) | P |
| SBP, *mmHg* | 131 ± 13.6 | -0.187<br>(-2.6; 2.2) | 131 ± 13.8 | 0.088<br>(-2.3; 2.5) | 0.275<br>(-3.2; 3.7) | 0.875 |
| DBP, *mmHg* | 75 ± 9.07 | -1.590<br>(-3.3;0.08) | 76 ± 9.14 | 0.064<br>(-1.6; 1.7) | 1.654<br>(-0.72; 4.0) | 0.172 |
| PP, *mmHg* | 108 ± 10.4 | -0.033<br>(-0.77; 0.70) | 108 ± 10.5 | 0.363<br>(-0.37; 1.1) | 0.396<br>(-0.65;1.4) | 0.455 |

SBP, systolic blood pressure; DBP, diastolic blood pressure; PP, pulse pressure (SBP-DBP). Data expressed as mean ± standard deviation or mean (95% Confidence Interval, CI). ANCOVA Model adjusted by sex, age, and sequence of treatments administration. No differences were observed.

**Supplementary Table 5.** Changes in lipid parameters at 4 weeks of interventions

| Intervention |  |  |  |  | Change inter-treatments |  |
| --- | --- | --- | --- | --- | --- | --- |
| Variable | OO |  | EVOO |  | EVOO *versus* OO |  |
|  | Post-int | Change | Post-int | Change | Mean (95%CI) | P |
| Cholesterol, *mg/dL* |  |  |  |  |  |  |
| Total | 203 ± 36 | -0.121<br>(-4.8; 4.5) | 206 ± 35 | 0.538<br>(-4.1; 5.2) | 0.659<br>(-7.2; 5.9) | 0.844 |
| HDL | 47.9 ± 11.6 | -0.220<br>(-1.7;1.3) | 47.5 ± 11.1 | -0.121<br>(-1.6; 1.4) | 0.099<br>(-2.0; 2.2) | 0.927 |
| LDL | 128 ± 30 | 1.47<br>(-3.2; 6.1) | 131 ± 34 | -0.570<br>(-5.2; 4.1) | -2.04<br>(-8.6;4.5) | 0.543 |
| Triglycerides, *mg/dL* | 136 ± 69 | -6.84<br>(-19;5.8) | 134 ± 62 | 6.14<br>(-6.5;19) | 13.0<br>(5.0;31) | 0.155 |
| Atherogenic ratios |  |  |  |  |  |  |
| Total Cholesterol/HDL | 4.42 ± 1.04 | 0.053<br>(-0.08;0.18) | 4.50 ± 1.03 | 0.085<br>(-0.05;0.22) | 0.032<br>(-0.15;0.22) | 0.736 |

**Suppl. Table 5 (cont)**
|  |  |  |  |  |  |  |
| --- | --- | --- | --- | --- | --- | --- |
| LDL/HDL | 2.79 ± 0.79 | 0.084<br>(-0.38;0.21) | 2.88 ± 0.89 | 0.050<br>(-0.07;0.17) | -0.034<br>(-0.21;0.14) | 0.697 |
| Triglycerides/HDL | 3.12 ± 2.01 | -0.154<br>(-0.51;0.20) | 3.06 ± 1.75 | 0.175<br>(-0.18;0.53) | 0.330<br>(-0.17;0.83) | 0.192 |

HDL, High density lipoproteins. LDL, low density lipoproteins. Data expressed as mean ± standard deviation or mean (95% Confidence Interval, CI). ANCOVA Model adjusted by sex, age, and sequence of treatments administration.

**Supplementary Table 6.** Changes in C reactive Protein, Thyrotropin, and 25-OH-Vitamin D at 4 weeks of interventions

| Intervention |  |  |  |  | Change inter-treatments |  |
| --- | --- | --- | --- | --- | --- | --- |
| Variable | OO |  | EVOO |  | EVOO *versus* OO |  |
|  | Post-int | Change | Post-int | Change | Mean (95%CI) | P |
| 25-OH-Vit D, *ng/mL* | 21.7 ± 7.27 | 0.705 | 21.9 ± 7.70 | 0.194 | -0.511 | 0.426 |
|  |  | (-0.19; 1.6) |  | (-0.70; 1.1) | (-1.8; 0.75) |  |
| TSH (log) | 0.25 ± 0.37 | -0.003 | 0.26 ± 0.35 | -0.015 | -0.012 | 0.676 |
|  |  | (-0.04;0.04) |  | (-0.05;0.02) | (-0.07; 0.04) |  |
| CRP | 0.60 ± 0.22 | -0.005 | 0.60 ± 0.21 | 0.24 | 0.029 | 0.345 |
|  |  | (-0.05; 0.04) |  | (-0.02; 0.07) | (-0.03;0.09) |  |
| hsCRP (log) | 0.57 ± 0.31 | 0.024 | 0.59 ± 0.30 | -0.007 | -0.031 | 0.391 |
|  |  | (-0.02;0.07) |  | (-0.06;0.04) | (-0.10;0.04) |  |

25-OH-VitD, 25-Hydroxy-Vitamin D; TSH, thyrotropin; CRP, C Reactive Protein; high sensitivity CRP. Data expressed as mean ± standard deviation or mean (95% Confidence Interval, CI). ANCOVA Model adjusted by sex, age, and sequence of treatments administration.

**Supplementary Table 7.** Changes in renal function related parameters at 4 weeks of interventions

| Intervention |  |  |  |  | Change inter-treatments |  |
| --- | --- | --- | --- | --- | --- | --- |
| Variable | OO |  | EVOO |  | EVOO *versus* OO |  |
|  | Post-int | Change | Post-int | Change | Mean (95%CI) | P |
| Urea, *mg/dL* | 36 ± 8.66 | 0.923<br>(-0.58; 2.4) | 35 ± 8.06 | 1.17<br>(-0.33; 2.7) | -0.253<br>(-2.4; 1.9) | 0.815 |
| Creatinine, *mg/dL* | 0.79 ± 0.19 | 0.024<br>(-0.004;0.05) | 0.78 ± 0.18 | 0.028<br>(-0.000;0.05) | 0.004<br>(-0.03; 0.04) | 0.842 |
| GFR, *mL/min/1.73m<sup>2</sup>* | 86 ± 8.69 | -1.47<br>(-3.4; 0.47) | 87 ± 8.18 | -1.46<br>(-3.4; 0.51) | 0.004<br>(-2.8; 2.8) | 0.998 |
| Uric acid, *mg/dL* | 5.86 ±1.49 | -0.147*<br>(-0.29; -0.05) | 5.81 ±1.39 | -0.125<br>(-0.27; 0.02) | 0.022<br>(-0.18; 0.22) | 0.830 |

GFR, glomerular filtration rate. Data expressed as mean ± standard deviation or mean (95% Confidence Interval, CI). ANCOVA
Model adjusted by sex, age, and sequence of treatments administration. \*p<0.05

